# Implementing an Electronic Health Record System in a Tertiary Care Surgical Oncology Setup – A Mixed Method Analysis

**DOI:** 10.1101/2025.07.12.25331172

**Authors:** Amila Prathibha Nellihela, Shamika Kavindi Gunaratne, Vasthsal Chinthaka Bandaranayake, Jayalthge Ruchika Nirmalie Senevirathne, Theekshana Pathirana, Malinga Gallala, Jayanjana Asanthi, Karunanithy Pirahanthan, Sulakshi Nikeshala Karunananayake, Abheetha Abegunasekara, Tharindu Saranga Jayasinghe, Mahesh Senarathne

**Author notes:** **Corresponding Author:** Amila Prathibha Nellihela, **Email**, **Contact**.

## Abstract

**Objective:** In Sri Lanka, resource limitations have led to the continued use of paper-based records for patient management. We implemented a cloud-based Electronic Health Record (EHR) system in a tertiary surgical oncology unit, running it alongside the existing paper system. The EHR provided authorised, real-time remote access to patient data, digital theatre scheduling, and facilitated multidisciplinary team collaboration.

**Methods:** Twenty-six healthcare workers (consultants, medical officers, nursing officers, trainees, and clerical staff) completed an online questionnaire assessing the EHR’s usability, user satisfaction, and impact on workflow. We prospectively tracked and compared key time metrics between the paper and EHR systems, including theatre list preparation times and cancer biopsy turnaround (biopsy-to-diagnosis interval), to evaluate efficiency gains.

**Results:** Most participants (84.6%) used the EHR routinely. Users rated the system as highly intuitive, user-friendly, easily accessible, and simple for data entry (mean ratings ∼ 4.0 out of 5). Overall satisfaction was high (mean 4.31/5), though system speed was rated slightly lower (mean 3.92), and technical glitches were noted (mean 3.65). Adequate training was associated with significantly higher satisfaction (p<0.05), and satisfaction correlated with perceived intuitiveness (r=0.43) and ease of use (r=0.60). The EHR reduced average theatre list preparation time from 4 minutes 6 seconds (paper) to 2 minutes 24 seconds, saving approximately 1 minute 42 seconds per list(p<0.001). Similarly, the median biopsy-to-diagnosis interval decreased from 14.95 days with the paper process to 8.40 days with the EHR’s notification system– an average reduction of 6.55 days(p<0.001).

**Conclusion:** Implementing a customised EHR system in a resource-limited surgical oncology setting significantly improved workflow efficiency, reduced diagnostic delays, and enhanced data accessibility and team coordination. Users reported high satisfaction, but challenges such as technical limitations, infrastructure issues, and resistance to change persist. Targeted training, supportive infrastructure, and stakeholder engagement are recommended to sustain the EHR integration and promote greater adoption.

**Highlights:**

- Electronic health records enhance workflow, data access, and team collaboration.
- Electronic health records significantly reduced biopsy-to-diagnosis delays.
- High user satisfaction is associated with intuitive design and adequate training.
- Technical issues and system speed were primary challenges for users.
- Targeted training and robust infrastructure are vital for successful implementation.

## Introduction

Electronic Health Record (EHR) systems facilitate real-time patient data management, improving documentation, patient safety, and multidisciplinary collaboration [1-3]. Despite demonstrated benefits globally, adoption in resource-limited settings remains challenging, hindered by infrastructure deficits and user resistance [4-6].

In Sri Lanka, government hospitals primarily rely on paper records due to fragmented policies and limited resources. Local initiatives, like the Hospital Health Information Management System (HHIMS), have demonstrated feasibility and benefits but face scale-up challenges due to interoperability issues [7-11]. This study addresses a gap by implementing and evaluating a specialised EHR in a tertiary surgical oncology unit, an underexplored context, aiming to provide evidence for broader adoption.

Electronic Health Record (EHR) systems are secure digital platforms that enable real-time access to and management of patient data (demographics, medical history, investigations, treatments, and follow-up care) [1]. Integrating EHRs into healthcare has been shown to improve documentation quality, patient safety, and multidisciplinary collaboration [2-3]. Globally, policies such as the U.S. HITECH Act and World Health Organisation eHealth initiatives have prioritised EHR adoption to strengthen health system efficiency and data-driven care coordination [4].

The concept of EHRs dates back to the 1960s, with the Mayo Clinic being one of the first to implement a hospital-wide EHR. Numerous studies have since demonstrated improvements in care quality and even cost reductions following EHR implementation, and international health agencies continue to advocate for EHR use to enhance quality of care, provider communication, and cost containment [1]. These anticipated benefits have driven various national strategies to accelerate EHR uptake, including incentive programs like the HITECH Act.

Despite well-documented benefits, implementing EHRs in real-world settings faces persistent challenges. Common barriers include user resistance, inadequate infrastructure, poor system interoperability, insufficient training and technical support for users, and low computer literacy among staff [5-6]. Resistance to changing longstanding paper-based workflows is frequently reported, as clinicians may hesitate to trust a new digital system with critical patient information. In resource-limited healthcare environments, high upfront costs and unreliable network connectivity can further hinder EHR adoption [6].

These challenges are especially pronounced in low- and middle-income countries such as Sri Lanka, where limited funding, fragmented national strategies, and minimal technical support have impeded digital health transitions [7-8]. Most government hospitals in Sri Lanka continue to rely on paper medical records. Nevertheless, there have been national efforts (e.g., the Hospital Health Information Management System [HHIMS] and the National Digital Health Guidelines and Standards [NDHGS]) aimed at expanding EHR use across public institutions [9]. To date, adoption of EHRs in the public sector has been piecemeal; there is no comprehensive national EHR system in operation—instead, various small-scale, often privately funded, EHR solutions function in isolated facilities. A national EHR platform remains under development, struggling with financial, infrastructural, and interoperability hurdles [10-11].

Previous local initiatives have nevertheless demonstrated the potential benefits of EHR adoption. For instance, a pilot project supported by the Austrian/Swiss Red Cross introduced EHRs in 20 hospitals in Sri Lanka’s Eastern Province, leading to faster record retrieval and improved clinical efficiency [12]. Similarly, an evaluation of the HHIMS platform in four Sri Lankan hospitals found high patient and staff satisfaction, with 85% of patients reporting improved quality of care [13]. Despite these efforts, the absence of a unified, nationwide EHR system continues to hinder the standardisation and scalability of digital health solutions. The Sri Lankan Ministry of Health has outlined plans for a National Electronic Health Record (NeHR) as part of a Digital Health Blueprint, aiming to establish lifelong electronic health records for all citizens [11]. However, progress has been slow, and success will depend on overcoming persistent challenges, particularly ensuring that different systems can interoperate effectively [14].

## Methods

### Study Design and Setting

This study used a convergent parallel mixed-methods design, collecting quantitative and qualitative data simultaneously to gain comprehensive insights into EHR implementation outcomes.

The study occurred at the Surgical Oncology Unit, Teaching Hospital Anuradhapura, Sri Lanka, one of the largest tertiary care centres in Sri Lanka, transitioning from a purely paper-based to an EHR-supported environment (Images 1 and 2).

Before this study, the unit managed patients with an entirely paper-based system: clinic visit notes were handwritten in patient-held record books, blood investigation results were transcribed into these books, and radiology/imaging and pathology reports were printed for patients to carry. This manual process often led to missing or damaged records, delays in retrieving past results, and repeated hospital visits for patients checking if results were ready.

To address these inefficiencies, we developed and implemented a tailored, modular cloud-based EHR system for the unit. Authorised users could access the system on hospital computers and remotely via a secure internet connection, enabling real-time updates and coordination across radiology, pathology, operating theatres, and clinics. The EHR was designed in accordance with Sri Lanka’s national eHealth guidelines [11] and was hosted on a secure Amazon Web Services cloud server with appropriate data encryption and backup protocols. The EHR was introduced while the paper record system continued in parallel, ensuring no disruption to patient care during the transition.

### EHR System Features

The pilot EHR included the following key modules and capabilities:

- **Patient Data Entry & Longitudinal Records:** Staff can record and update patient demographics, medical history, examination findings, investigation results, operative notes, discharge summaries, and follow-up clinic notes. Patients are identified using multiple parameters (e.g., hospital ID, clinic number, national ID) and are issued barcode-enabled identity cards to facilitate quick retrieval of digital records during clinic visits.
- **Theatre List Management:** An interactive scheduling interface allows creation and editing of surgical theatre lists, replacing the previous handwritten lists. Theatre schedules can be shared electronically with anesthesiology and nursing teams or printed as needed, improving efficiency and consistency.
- **Advanced Search & Data Export:** Users can query the database using flexible filters (e.g., by diagnosis, date range) and export data in multiple formats (PDF, Excel, CSV). This function facilitates clinical audits and research by enabling structured data extraction compatible with statistical software.
- **Administrative Control Panel:** An admin dashboard supports user account management, role-based access control, and system usage monitoring. Administrative privileges are restricted to designated staff. All user actions are logged for auditability, enhancing accountability and security.
- **Multidisciplinary Team (MDT) Platform:** This module enables clinicians to submit oncology cases for multidisciplinary tumour board discussions. MDT members can concurrently review patient records, imaging, and pathology reports during meetings and record the team’s recommendations. After each meeting, the system generates a summary of the MDT decisions (including attendee list and agreed management plans), which can be printed for the patient’s file to improve care continuity.
- **Patient Notification System:** The EHR integrates with an SMS/email gateway to notify patients and clinicians of essential updates automatically. For example, when a pathology report is finalised, the responsible surgical oncology staff receive an alert and an SMS is simultaneously sent to the patient to collect the results or attend for follow-up. The system also sends reminders for scheduled surgeries and notifications of any postponements, reducing unnecessary hospital visits and ensuring timely follow-ups.
- **Data Security & Compliance:** The platform employs multiple security layers, including role-based user access (each user has a unique login with permissions according to their role), end-to-end encryption (SSL) for data transmission, and weekly automated backups with disaster recovery support. A formal non-disclosure agreement with the software developer was in place to maintain confidentiality. All measures align with Sri Lanka’s National Digital Health Guidelines and Standards to protect patient data privacy and ensure system integrity.

### Study Population and Sampling

The EHR system was launched unit-wide across the surgical oncology outpatient clinic, inpatient ward, and operating theatre. For the quantitative evaluation of system impact, we prospectively tracked **100 consecutive patients** with histologically confirmed malignancies who entered the unit’s care after EHR implementation. These patients had their clinical information recorded in both the traditional paper record and the EHR from the point of first visit onward, allowing paired comparisons of process times (paper vs. electronic) for those patients.

For the user experience assessment, we invited all healthcare professionals in the unit who had at least one month of experience using the new EHR system. This included 26 staff members (8 consultant surgeons, 10 medical officers, six nursing officers, one postgraduate trainee, and 1 data entry officer), and all 26 agreed to participate by completing the survey.

### Data Collection and Outcomes

We defined two primary outcomes to assess the impact of EHR implementation:

1. **Workflow efficiency:** We measured the time required for two key processes under the paper-based versus EHR system: (a) preparation of the daily surgical theatre list, and (b) the interval from performing a cancer biopsy to the availability of the pathology report (biopsy turnaround time). We recorded and averaged the times for each process using both methods and calculated the time saved with the EHR for each.
2. **User perceptions and satisfaction:** Using an anonymous online questionnaire, we collected feedback on multiple aspects of the EHR’s usability and usefulness. Survey domains included system intuitiveness, ease of data entry, data accessibility, interface design, system speed, adequacy of training, and quality of technical support. Responses were given on 5-point Likert scales (1 = strongly disagree/very poor, 5 = strongly agree/excellent), along with categorical options for training adequacy and free-text fields for additional comments.

Basic demographic information about the survey respondents (professional role and duration of EHR use) was also gathered. The Likert-scale ratings were analyzed statistically: a one-way ANOVA was used to test for differences in overall satisfaction between groups who felt they had adequate training versus those who did not, and Pearson correlation coefficients were calculated to examine relationships between overall satisfaction and specific usability factors (e.g., correlation between satisfaction and perceived intuitiveness). For the efficiency outcomes, we used paired *t*-tests to compare mean times for the theatre list preparation and biopsy turnaround under the two systems. A p-value < 0.05 was considered statistically significant for all analyses. The study received ethical approval from the institutional review board, and all participating staff provided informed consent for the survey.

### Ethical Approval

Obtained from the Ethics Review Committee, Faculty of Medicine, Rajarata University (Approval No. ERC/2022/32). Written informed consent was obtained from all participants. Data were anonymised and securely stored.

## Results

### User Satisfaction and Usability

A total of 26 healthcare workers completed the post-implementation survey. Of these, 84.6% had been using the EHR system regularly for six months at the time of evaluation. Overall feedback on the EHR was positive. Users rated the system as intuitive and easy to learn (mean score ∼4.1 out of 5), with a similarly high rating for interface friendliness (mean ∼4.1). Accessing patient information was considered straightforward (mean ∼4.1), and users felt the EHR fulfilled clinical data management needs effectively (mean 4.04). Data entry was also well-rated (mean 4.00), indicating that staff found it reasonably easy to input and update patient information electronically. In terms of support, the responsiveness of technical support services received one of the highest scores (mean 4.54), reflecting that users felt well-supported by IT staff during the rollout. These usability findings are summarised in Figure 1.

**Figure 1.**
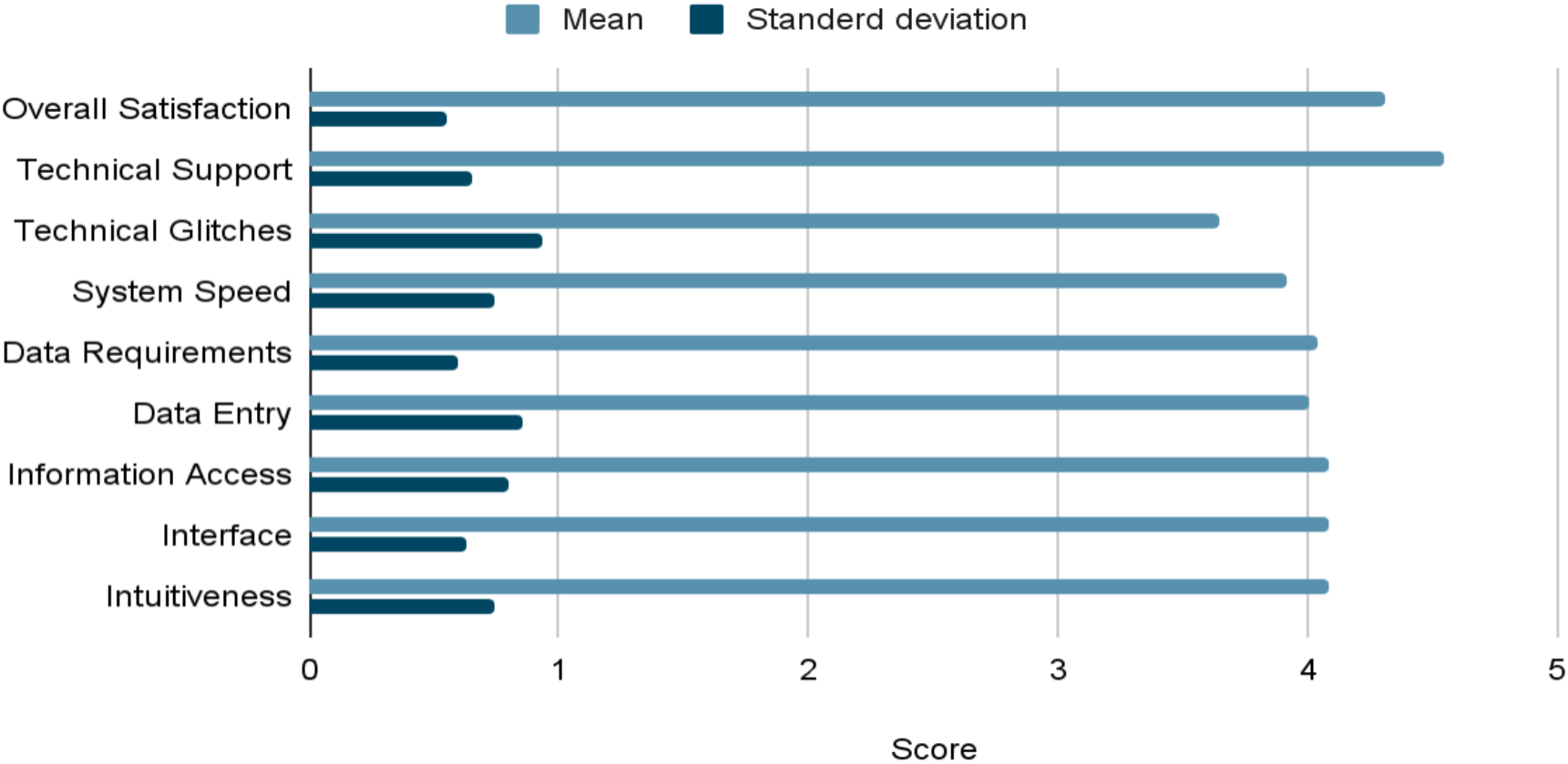
Descriptive statistics of Usability and User Experience of the EHRs

Despite the generally high usability scores, a few concerns emerged. System speed was rated slightly lower (mean 3.92), suggesting that some users experienced occasional slowness or lag. Technical glitches were the most notable issue, with a lower mean score of 3.65 indicating that users encountered software errors or downtime relatively frequently (Figure 1). Opinions on training were mixed: 34.6% of users felt the training provided was only “somewhat adequate,” and another 30% rated it “adequate,” implying a substantial proportion saw room for improvement in training.

Notably, users who reported having received “adequate” training showed higher overall satisfaction with the EHR (average satisfaction score 4.47) compared to those who felt training was only “somewhat adequate” (average 4.00). This difference in satisfaction between training groups was statistically significant (one-way ANOVA, F = 5.016; p < 0.05). There were also significant positive correlations between overall satisfaction and specific usability aspects. In particular, overall satisfaction was moderately correlated with perceived system intuitiveness (r = 0.43, p < 0.05) and interface friendliness (r = 0.39, p < 0.05). The strongest association was between satisfaction and the ease of entering/updating patient information (r = 0.60, p < 0.01). These correlations between satisfaction aspects are depicted in Figure 2. These findings suggest that improving the training process and focusing on user-centred design elements could directly enhance user satisfaction with the EHR.

**Figure 2.**
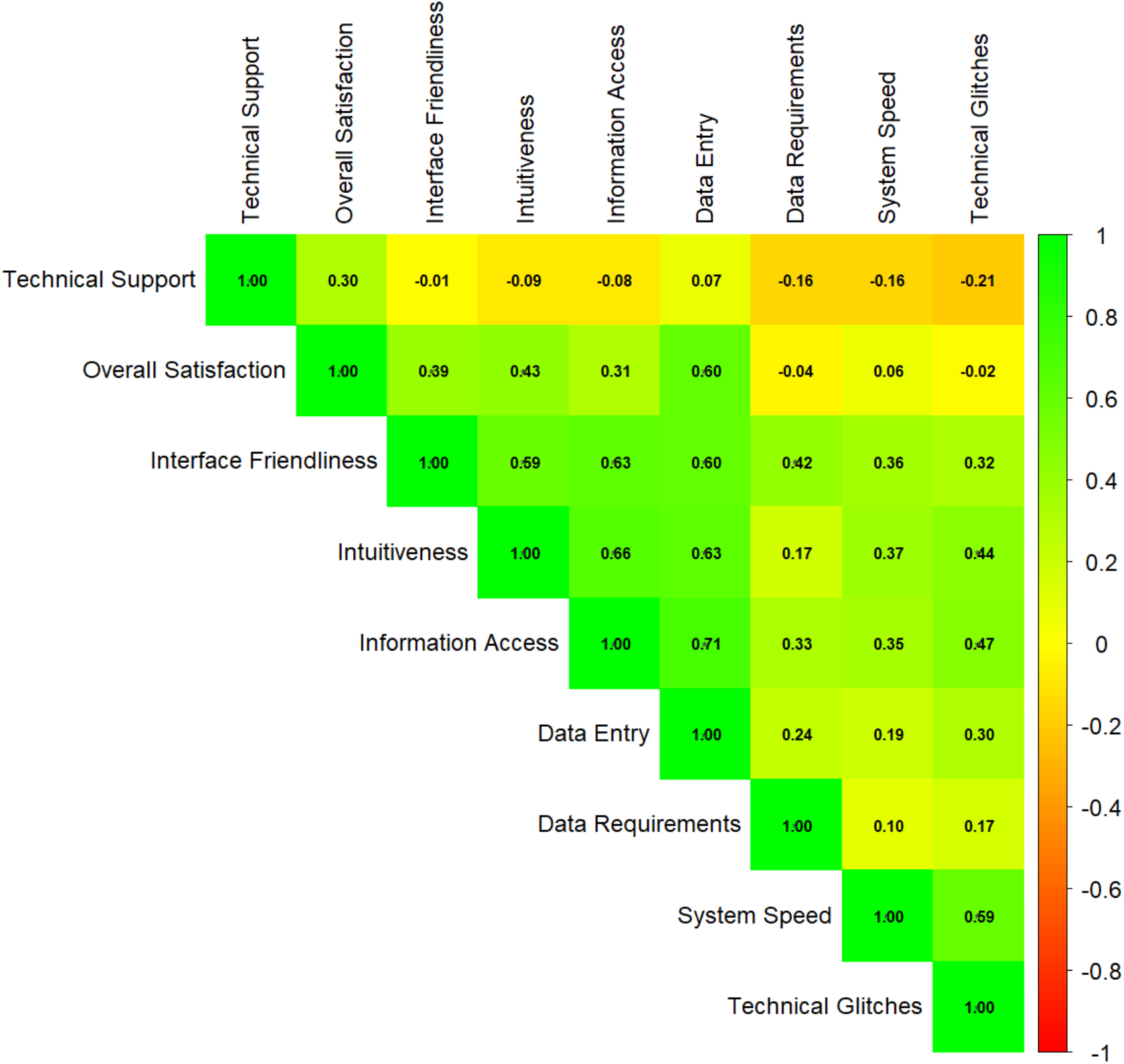
Correlation plot of Usability and User Experience of the EHRs

### Workflow Efficiency Outcomes

The introduction of the EHR significantly improved efficiency in the unit’s workflow. Preparing the daily operating theatre list – a task that was previously done by hand – became much faster with the EHR. The average time to compile a theatre list manually was **4 minutes 6 seconds**, whereas using the EHR’s digital scheduling module, it averaged **2 minutes 24 seconds**. This difference of roughly 1 minute 42 seconds represents a **40% reduction** in list preparation time with the electronic system, a change that was statistically highly significant (Table 1; paired *t*-test, t = 12.904, p < 0.0001). Throughout many surgical lists, this time savings can translate into substantially reduced administrative burden and quicker communication of the schedule to the surgical team.

**Table 1:** Efficiency Gains in Time-Sensitive Processes.

| Task | Handwritten<br>Time | EHR Time | Time Saved | t-<br>value | Statistical<br>Significance |
| --- | --- | --- | --- | --- | --- |
| Theater List<br>Creation | 4 min 6 sec<br>(SD = 48s) | 2 min 24<br>sec (SD =<br>25s) | 1 min 42 sec | 12.90<br>4 | $p < 0.0001$ |
| Biopsy to Cancer<br>Diagnosis | 14.95 days<br>(SD = 3.55) | 8.40 days<br>(SD = 2.91) | 6.55 days | 6.402 | $p < 0.0001$ |

Similarly, implementing the EHR dramatically shortened the biopsy result turnaround time. Under the paper-based process, patients waited an average of **14.95 days** from the date of biopsy to receive their pathology report. After the EHR introduced automated notifications and tracking, the average biopsy-to-diagnosis interval decreased to **8.40 days**. This is an improvement of **6.55 days** on average, meaning patients learned of their diagnoses nearly a week sooner with the EHR-supported workflow. This reduction in diagnostic delay was also statistically significant (Table 1; t = 6.402, p < 0.0001). Accelerating the delivery of biopsy results allows for earlier oncology treatment planning and may improve patient anxiety and outcomes. It is also noteworthy that running the EHR in parallel with the paper system did not result in any loss of information; on the contrary, it provided a redundancy that ensured patient data were captured accurately in both formats during the transition phase. Visual comparisons between handwritten and EHR-generated theatre lists are presented in Image 1 and Image 2, respectively.

**Image 1:**
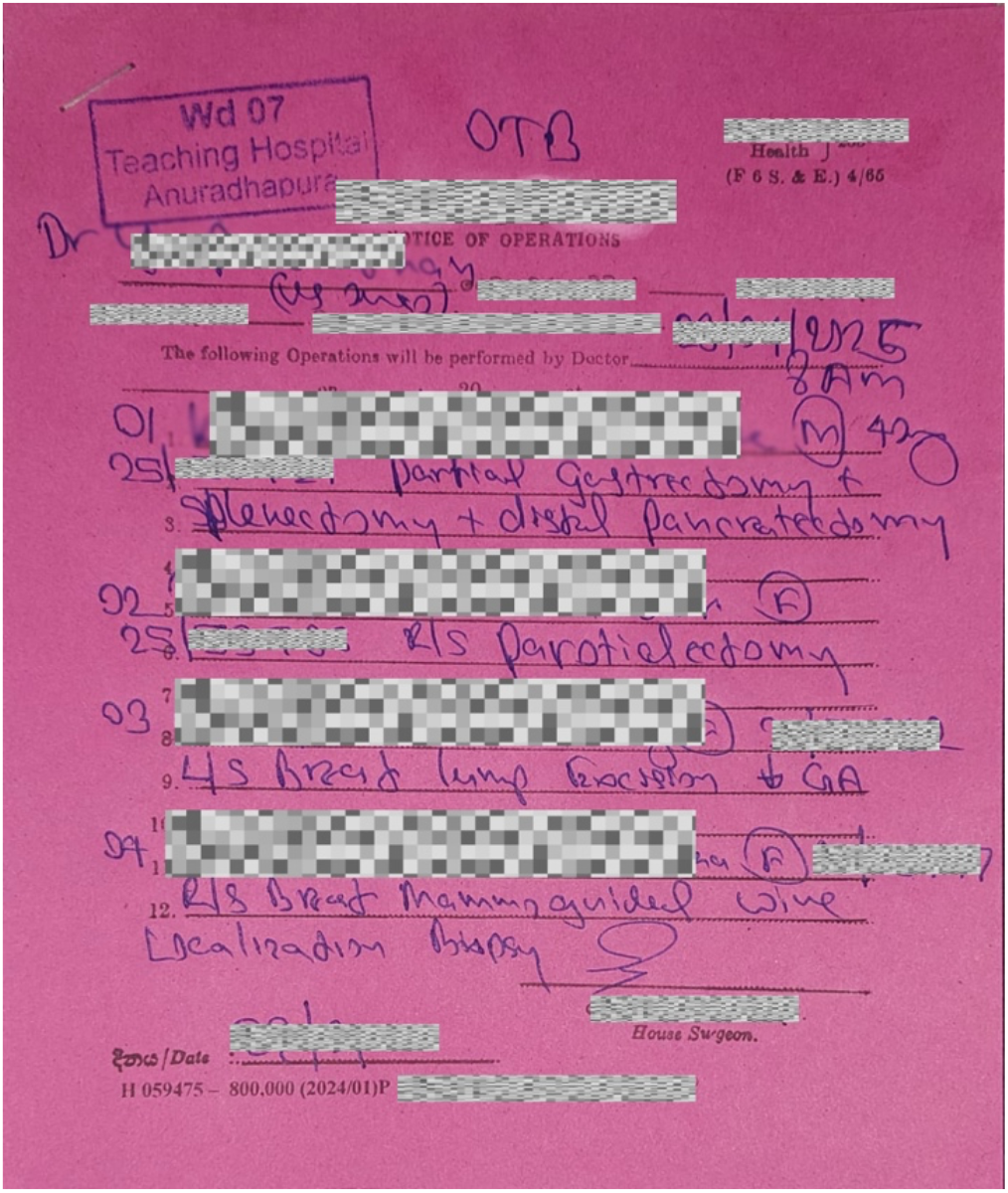
Handwritten theatre list

**Image 2:**
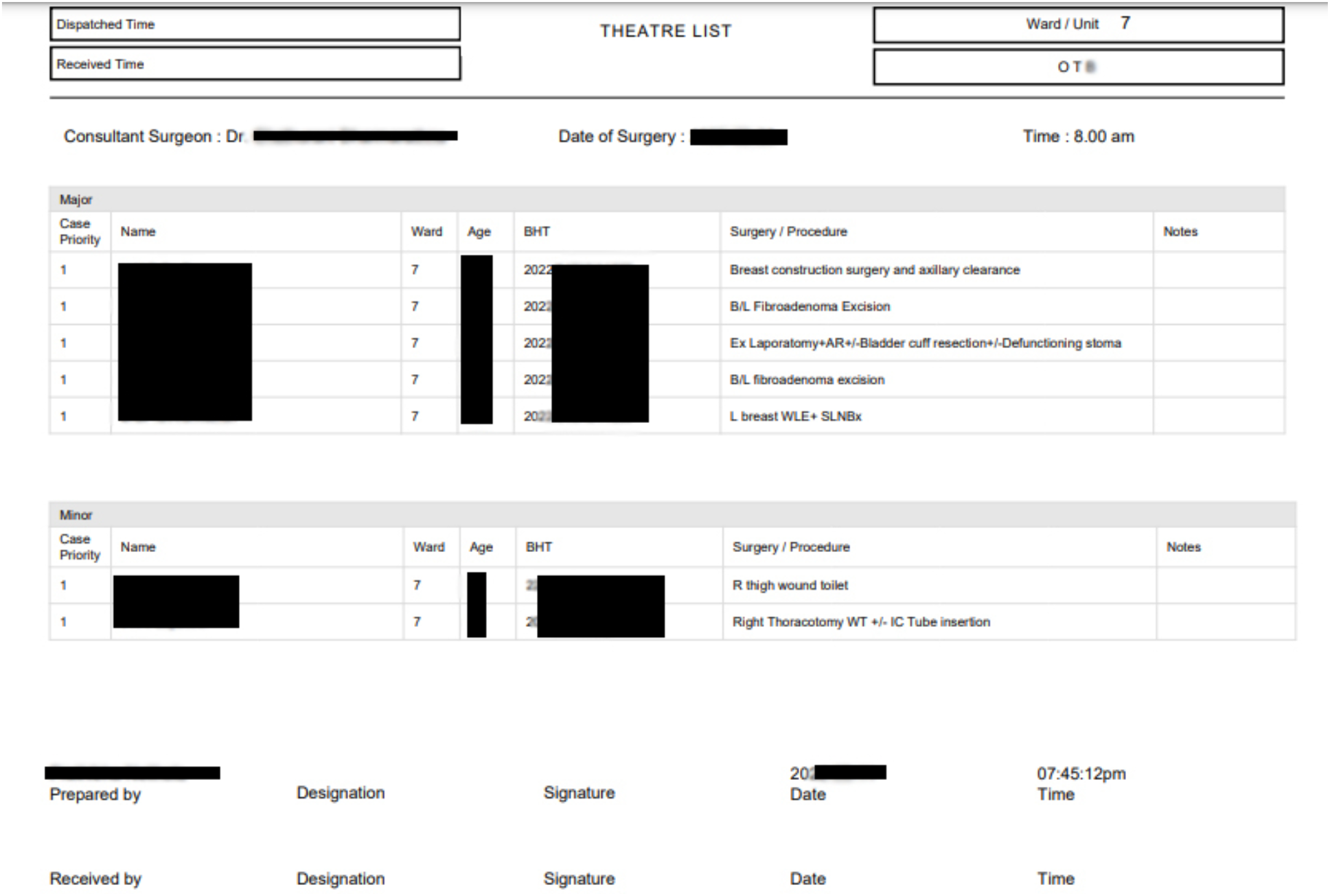
EHRs Generated Theatre List

## Discussion

This mixed-method pilot study demonstrates that implementing a cloud-based EHR system in a resource-limited, high-volume surgical oncology unit can yield substantial benefits. We observed marked improvements in clinical workflow efficiency and significant reductions in critical time intervals (e.g., surgical scheduling and diagnostic turnaround times) after the EHR implementation. These quantitative gains align with global evidence that EHR systems streamline healthcare processes and improve care timeliness [1-3]. Importantly, the new system was well-received by end-users, indicating that even in a low-resource setting, a carefully tailored EHR can achieve high user acceptance and usability.

User satisfaction in our study was notably high, particularly regarding the EHR’s intuitiveness, user-friendly interface, and data accessibility. These positive perceptions mirror outcomes from other Sri Lankan eHealth initiatives; for example, the HHIMS electronic records project also reported improved quality of care and streamlined provider workflows with digital systems [12-13]. Our findings underscore the critical role of adequate training: staff who felt properly trained were significantly more satisfied with the EHR, reinforcing global evidence on continuous training and user support as keys to successful EHR adoption [5-6].

Several challenges, however, tempered enthusiasm for the new system. Users frequently cited slow system speeds and minor technical glitches as irritants—issues commonly reported in EHR implementations, particularly in low-resource settings [6,8]. Despite the cloud-based architecture, hospital internet connectivity occasionally disrupts system performance, highlighting the importance of concurrent investments in robust IT infrastructure (e.g., reliable network bandwidth, updated hardware) to support digital health solutions effectively.

Resistance to change emerged as another significant barrier, particularly among nursing staff who perceived electronic documentation as increasing their workload. This emphasises the need for careful workflow redesign and proactive stakeholder engagement. Involving end-users early in the EHR development process, providing role-specific training, and establishing “clinical champions” can help overcome resistance and reinforce perceived system benefits [2]. As familiarity with the EHR grows and its time-saving advantages become apparent, resistance is expected to diminish.

Concerns around data privacy and patient confidentiality were also noted, consistent with global apprehensions during healthcare digitisation [14]. These were effectively managed through rigorous adherence to national digital health standards, employing strict security features such as encrypted logins, audit trails, and role-based access controls. These safeguards likely contributed to overall acceptance, reassuring users and patients about information security.

From a broader perspective, our experience demonstrates the feasibility and value of deploying a purpose-built EHR system at an institutional level, especially when a comprehensive national EHR system is not yet available. Currently, Sri Lanka’s digital health efforts remain fragmented and institution-driven, although a national EHR framework is under development [11]. Interim, locally developed EHR solutions that comply with national data standards and emphasise interoperability can effectively bridge existing gaps. Such modular systems hold the potential to integrate into broader national platforms once established.

This study significantly contributes to the growing evidence base supporting digital health adoption in low- and middle-income countries. It provides practical insights into the technical, behavioural, and organisational enablers and barriers to EHR implementation. However, the single-centre pilot nature and relatively short observation period limit generalizability. Future research should encompass multi-centre studies, longer-term evaluations of clinical outcomes, cost-effectiveness, and patient satisfaction to more comprehensively assess EHR impacts within Sri Lanka’s healthcare system. Investigating methods for scaling and sustaining EHR implementations, as well as interfacing these systems with evolving national eHealth policies, will be particularly beneficial.

## Conclusion

In summary, implementing a tailored, cloud-based EHR system in a tertiary surgical oncology unit in Sri Lanka significantly improved workflow efficiency, reduced diagnostic delays, and received positive feedback from clinical staff. The system demonstrated clear benefits in reducing administrative delays (such as scheduling and diagnostic reporting times) and enhancing clinical data accessibility, underlining the potential of thoughtfully designed health IT interventions in resource-limited environments.

Persistent challenges, including technical limitations, infrastructure deficiencies, and initial user resistance, must be proactively managed through continuous training, reliable technical support, and robust stakeholder involvement. In the absence of an immediate nationwide EHR solution, locally developed systems that adhere to national standards and emphasise interoperability represent effective interim measures for digitising patient records. Our pilot implementation offers valuable, context-specific insights and serves as proof-of-concept to inform broader digital health initiatives in Sri Lanka and similar low-resource healthcare settings.

## Data Availability

All data produced in the present work are contained in the manuscript

## Conflicting interests

The authors declare that there are no conflicts of interest.

## Funding

This research received no specific grant from any funding agency in the public, commercial, or not-for-profit sectors and was personally funded by the principal author.

## Ethical approval

Ethical approval for this study was granted by the Ethics Review Committee, Faculty of Medicine, Rajarata University (Approval No. ERC/2022/32). Written informed consent was obtained from all participants.

## Guarantor

Amila Prathibha Nellihela acts as the guarantor, accepting full responsibility for the conduct of the study, access to the data, and decision to publish.

## Contributorship

**Amila Prathibha Nellihela**: Conceptualisation, Methodology, Formal Analysis, Investigation, Project Administration, Supervision, Writing – Original Draft, Supervision, Writing – Review & Editing.

**Shamika Kavindi Gunaratne**: Co-Principal Author – Conceptualisation, Methodology, Formal Analysis, Investigation, Project Administration, Writing – Original Draft, Writing – Review & Editing.

**Jayalthge Ruchika Nirmalie Senevirathne**: Conceptualization, Methodology, Formal Analysis, Investigation, Project Administration, Writing – Original Draft, Writing – Review & Editing.

**Vathsal Chinthaka Bandara** Supervision, Methodology, Validation, Writing – Review & Editing.

**Theekshana Pathirana**: Supervision, Methodology, Validation, Writing – Review & Editing.

**Malinga. Gallala**: Supervision, Methodology, Validation, Writing – Review & Editing.

**Jayanjana Asanthi (JA)**: Supervision, Methodology, Validation, Writing – Review & Editing.

**Karunanithy Pirahanthan**: Data Collection, Investigation, Writing – Review & Editing.

**Sulakshi Karunanayake**: Data Collection, Investigation, Writing – Review & Editing.

**A bheetha Abegunasekara**: Data Collection, Investigation, Writing – Review & Editing

**Saranga Jayasinghe**: Data Collection, Investigation, Writing – Review & Editing.

**Mahesh Senarathne**: Statistical Analysis, Validation, Writing – Review & Editing.

## Acknowledgement

We would like to sincerely acknowledge M. Rathnayake, R.M.M. Rathnayake, A.P.G.A. Indrajith, R.P.R.V. Pathirana, Y.A.S.D. Ranaweera, R.M.N.D. Rathnayake and K.N.O. Hettige for their invaluable contributions to the development of the Electronic Health Record system used in this study.

## Notes

### Competing Interest Statement

The authors have declared no competing interest.

### Funding Statement

This study did not receive any funding and was personally funded by the principal investigator

